# Supporting people to access social security payments through the ‘Special Rules for End of Life’: a qualitative study of the perspectives of patients, carers and health care professionals

**DOI:** 10.64898/2026.06.12.26355509

**Authors:** Joanna M Davies, Steve Marshall, Jamilla Hussain, Melanie Diggle, Maddy French, Juliet Stone, Geoff Fimister, Margaret Ogden, Katherine E Sleeman, Andy Bradshaw, Richard Harding

**Author notes:** **Correspondence to:** Cicely Saunders Institute of Palliative Care, Policy and Rehabilitation, Florence Nightingale Faculty of Nursing, Midwifery & Palliative Care, King’s College London, Bessemer Road, London SE5 9PJ, UK. joint senior authors (Andy Bradshaw, Richard Harding).

## Abstract

**Background:** People living with terminal illness face a double financial burden from additional costs and loss of earning for themselves and their carers. Social security benefits are intended to help alleviate some of this financial pressure, and in the UK and other countries people are eligible for fast-tracked access to financial support via the ‘Special Rules for End of Life’. One in 3 people who are eligible miss out on this support, yet there is limited evidence on the reasons for this take-up deficit.

**Objectives:** The aim of this study is to understand the barriers and facilitators to claiming benefits for terminally ill people from the perspectives of patients, carers, and health care professionals.

**Methods:** This is a qualitative study combining i) focus groups with healthcare professionals recruited via professional networks and social media, and ii) interviews with patients and carers recruited in hospital and hospice settings. We analysed the data using Practical Thematic Analysis

**Results:** Fifty-five multidisciplinary healthcare professionals participated in 11 focus groups, and we interviewed 10 patients and carers. We constructed five descriptive themes to summarise the data: Navigating priorities and uncertainty; positive impacts alongside a sense of shame and stigma; talking about money, difficulties and dividends; everybody’s – yet nobody’s – responsibility; and sticking points in the system.

**Conclusion:** The themes reveal several challenges that may contribute to people not taking up this financial support. However, discussions about access to benefits were also seen as a core part of holistic care, a positive way to offer support and a gateway to other discussions about end-of-life care preferences and decisions. Recommendations for policy and practice include evaluating the adoption of a diagnostic rather than a prognostic eligibility criteria, integrating discussions about benefits into existing processes such as advance care planning, and improving education and support for clinicians.

## Introduction

People living with terminal illness and their family carers often experience financial difficulties due to loss of earnings and the additional costs associated with serious illness. These costs can include out of pocket expenses for travel, medications, nutrition, equipment, heating, and home adaptations.^1–8^ Households living on a low income can spend as much as 98% of their income on these additional costs, with some decedents accumulating a ‘debt legacy’.^9,10^

Poverty and financial stress form part of the social dimension of the ‘total pain’ construct in palliative care, and can impact the emotional and psychosocial wellbeing of patients and families living with terminal illness.^11–13^ Financial hardship is also associated with worse physical symptoms including pain,^14^ and breathlessness^15^, and with patterns of service use including higher use of unplanned hospital care^16^ and less access to specialist palliative care in the last months of life.^17^ In several countries, including the UK,^18^ United States,^19^ Canada,^20^ and Australia,^21^ people living with a terminal illness are provided fast-tracked access to government financial support (benefits) to help alleviate some of the additional costs they face, and support independence and dignity as their illness progresses.

In England, Wales and Northern Ireland, under the ‘Special Rules for End of Life’ people in their last year of life (following an extension from a 61Zmonth prognosis, implemented incrementally across different benefits between 2022–2023) are eligible for fast-tracked access to benefits. This includes the non-means-tested disability benefits Personal Independence Payment (PIP) for working-age people (currently (2026-7) worth up to £194.60 per week depending on mobility needs), and Attendance Allowance (AA) for people of state pension age and above (currently worth up to £114.60 per week). For patients to access these benefits via the ‘Special Rules’ a doctor or a specialist nurse must submit information about their patient’s progressive, incurable disease via an SR1 form. This expedites the benefit application process without the need for further medical assessments.^18^

Recent analysis in England and Wales found that one in three people who die from an expected cause of death do not claim the non-means-tested benefits to which they are entitled under the ‘Special Rules’.^22^ This take-up deficit undermines the effectiveness of social security programmes and may increase costs in the long-term if people are less able to support their own care in the community. There are many reasons why people may not take-up the benefits they are entitled to, including barriers or lack of support in the system, previously rejected claims, stigma, or perceived ineligibility.^10, 23, 24^ Healthcare professionals can find discussing financial issues with patients challenging or may not recognise their role in helping to facilitate access to financial support.^25, 26^

To our knowledge, no one to date has studied the experience of claiming benefits under the ‘Special Rules for End of Life’. Therefore, the aim of this study is to understand the barriers and facilitators to claiming benefits for terminally ill people from the perspectives of patients, carers, and health care professionals. The findings are intended to inform the development of resources for healthcare professionals and to make policy recommendations to improve benefit take-up.

## Study design and methods

### Methodology

This was an exploratory qualitative study grounded in social constructivism (i.e., an epistemology that accepts multiple realities exist and meaning is contextual and situated).^27^ This approach recognises that research data is constructed through interactions between researchers and participants and that the researcher plays an active role in constructing and interpreting the meaning of data.^28, 29^

### Settings, participants and recruitment

Patients and carers were recruited from two acute hospitals (one in London and one in West Yorkshire) and two hospices (one in Wales and one in the West Midlands). Patients were eligible to participate if they were known to their clinical team to have a life-limiting illness with a prognosis of 12 months of less (or be the family member of someone with a life-limiting illness), were aged 18 years or older and able to participate in an interview lasting approximately one hour. Both those with and without experience of claiming benefits were eligible and patients and carer dyads could be interviewed separately or together.

Healthcare professionals were recruited through social media and professional networks, and we sought people who had experience working with patients and families living with life-limiting illness who were in the last year of life.

Maximum variation sampling,^30^ a purposive sampling frame, guided participant recruitment, aiming to capture diversity in patient and carer characteristics (including age, gender, ethnicity, and diagnosis) to inform understandings from diverse perspectives. For healthcare professionals we aimed to recruit a range of professions and specialties. The decision to stop recruiting for the focus groups was informed by information power i.e., how much information we had gathered and whether it was sufficient quality and depth for addressing our aim. For the interviews recruitment was curtailed by reaching resource (time and money) limits.^31^ Participants were not paid for their time.

### Patient and Public Involvement (PPI)

Three members of the public were involved in this work from the inception of the project and throughout. One PPI member (MO) was a co-applicant helping to plan this study and is a co-author on this paper. The PPI members met with the core study team (JD, SM and MD) three times throughout the study and helped to inform the recruitment strategy and topic guides, for example suggesting prompts and follow up questions.

### Data collection

This study involved semi-structured in-depth qualitative interviews with patients and carers and focus groups with healthcare professionals. We took a pluralistic approach to data collection integrating these different forms of data to provide complementary insight into the research topic, including the perspectives of healthcare professionals and patients and families.^32^

Interviews with patients and carers were conducted face-to-face, by telephone or online (Microsoft Teams) depending on participants’ preference. Each participant was interviewed once. The topic guide informed by the project advisory group and PPI members was developed and guided discussions around the experience of claiming benefits (see supplementary file p2-3).

Focus groups with healthcare professionals were organised pragmatically around availability and were both mixed and single specialty. A focus group topic guide was developed with input from the project advisory group and PPI members and focused on knowledge of the ‘Special Rules’, discussing benefits with patients, and completing applications (supplementary file p4-5).

The interviews and focus groups were carried out by SM, a specialist palliative care social worker and researcher with professional connections to some of the focus group participants. Recordings were auto transcribed verbatim using software and then checked and corrected (by SM and MD) before being anonymised.

### Ethics

Ethical approval for the study was granted in April 2024 by the NHS Health Research Authority (Dulwich Research Ethics Committee – Reference: 24/LO/0211). Informed consent was taken from all participants prior to data collection.

### Data analysis

Peer debriefing and discussion between SM and JD took place throughout data collection and included reflection on the construction of initial themes. The analysis was carried out by JD (a social epidemiologist) following a Practical Thematic Analysis approach using three steps of analysis: reading, coding and theming.^33^ Conceptually simpler than Reflexive Thematic Analysis, Practical Thematic Analysis is well-suited to applied health research studies being conducted by multi-disciplinary teams. This approach was well aligned to answering the research questions which have a strong practical and policy focus and a descriptive aim. JD read all the manuscripts in full and developed initial codes providing explicit descriptions of the content and then constructed themes to unite the codes and address the aims of the study. Data was coded in Microsoft Excel. The write-up of findings was treated as part of the analytic process, with themes being iteratively refined through critical reflecting on how best to tell the story of study findings comprehensively.^34^ Throughout analysis and write-up, co-authors acted as ‘critical friends’, ^35^ offering alternative interpretations and explanations of data, alongside supporting JD in engaging in introspective forms of reflexivity^36^ (i.e., critically examining how her own background and biases may have shaped the analysis and interpretation of findings).

## Results

### Sample characteristics

Ten patient and carer participants took place in 9 interviews (1 interview had a patient and carer present). Participants were recruited from hospital (3 in London, 3 in West Yorkshire) and hospice settings (1 in Wales and 3 in West Midlands). Patients had a range of diagnoses including cancer, heart failure, respiratory disease, liver disease and multiple long-term conditions. Four participants were female and 6 were male, most were of pension age with one working age participant. Seven participants were White British, two were British Asian and one had White Other ethnicity, all participants spoke English and all had experience of claiming benefits.

Fifty-five healthcare professionals participated in 11 focus groups, including 16 social workers, 12 benefits advisors, 10 clinical nurse specialists, 10 doctors, 5 therapists and 2 healthcare assistants. Participants were located across England and Wales and worked in specialist palliative care and in allied specialities such as cystic fibrosis and lung cancer care.

Five themes were constructed providing insight into the barriers and facilitators that healthcare professionals and patient’s experience in relation to the ‘Special Rules’ (figure 1). The supplementary file contains a list of initial codes (table 1) and additional supporting quotes for each of the themes.

**Figure 1:**
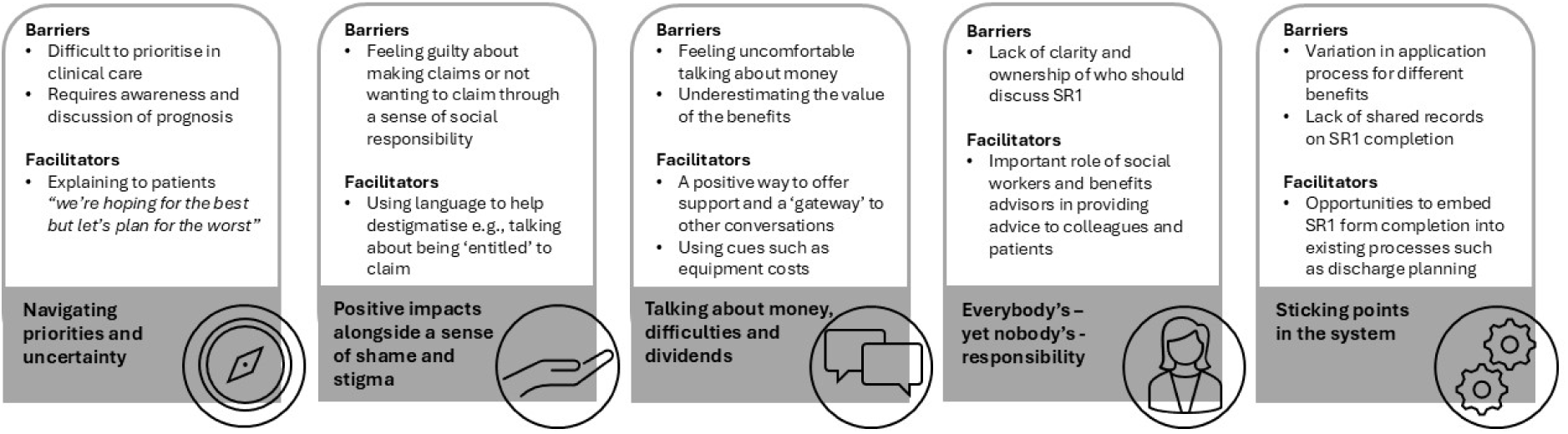
A summary of themes on barriers and facilitators to making benefit claims via the ‘Special Rules for End of Life’

### Themes

#### 1. Navigating priorities and uncertainty

Clinicians and social workers spoke about the practical challenges of completing an SR1 form (the form submitted by clinicians to confirm a patient is eligible for access to benefits via the ‘Special Rules’) when patients are very unwell, confused or unconscious. It can also be challenging to know whether a claim is worthwhile submitting if the patient is close to death, unlikely to be discharged home or is likely to be discharged to a local authority funded care home (people who enter state-funded residential or nursing care, or who have a hospital admission lasting 28 days or more, may have their disability benefit payments suspended or stopped in line with payment rules). Several doctors and nurses said that discussing benefits was often not a priority for them, or for the patient, when the patient is very unwell or during a busy clinic.

> *“If I’m going to be really honest, it’s not always my first thought because our patients are often acutely unwell, so their finances, while you know they are really important, in fact is not their first thought while they’re in hospital, so unwell.”* (nurse 9)

> *“In clinic is even harder, you’ve got 10 minutes, 15 minutes max…and in that 15 minutes you’re trying to tell them what they want to know clinically…sometimes you feel like you don’t want to open Pandora’s box.”* (doctor 9)

Healthcare professionals described how discussions about access to benefits under the ‘Special Rules’ are tied to discussions about limited prognosis, and this can make it challenging to discuss if the patients are not aware of, or have indicated that they do not want to discuss, their prognosis.

> *“They may not have already had that conversation with the patient about prognosis so, there’s almost like a fear of upsetting the patient by having these conversations [about benefits].”* (social worker 7)

Several social workers talked about the ethical dilemma of processing applications (SR1 forms) for patients who may not be fully aware of their prognosis.

> *“The patient wasn’t explicitly told exactly what this SR1 was…It was a bit of an ethical dilemma for myself…[but] I think it was the right thing for that patient. It would have caused a lot of, well, just another difficult conversation that she probably didn’t actually need at that point in time.”* (social worker 11)

Others spoke about how they reconciled completing an SR1 form with uncertainty about prognosis when speaking to patients.

> *“When I’m talking to the patient or the family about the SR1 form, I will sort of say to them ‘I really don’t know what’s going to happen within the next 12 months, I feel that you’d really benefit from having these benefits’…I’m trying to be a bit candid…I don’t want them to be told ‘this form says that you’re going to die within the next 12 months’ and I’ve not said that to them. So, I’ve said that to a few patients recently and they’ve been quite happy with it.”* (doctor 7)

> *“When you fill in forms and there’s a disconnect between what you’re writing and what you’re saying, it’s you know, we’re hoping for the best but let’s plan for the worst.”* (doctor 8)

Healthcare professionals reflected that discussions with patients and colleagues about access to benefits under the ‘Special Rules’ can be harder when prognosis is less clear, including with non-cancer conditions and some cancers such as haematological cancers. This was cited as a major barrier to completing SR1 forms. Healthcare professionals also spoke about a lack of advisory services for people with non-cancer conditions.

> *“But if someone has a blood cancer like a myeloma…we find it very difficult to get an SR1 …there’s a reluctance with certain cancers where, you know, the prognosis again is not predictable…and again, you know, infections, anything could happen in a year, but there’s a difficulty sometimes with getting those.”* (social worker 13)

> *“We have had problems over the years with getting advice to people without malignancy…The [cancer patient support centre name redacted]… don’t help with patients without malignancy…[I’ve] got quite frustrated that patients without malignancy…haven’t had that service available to them in the same way.”* (doctor 7)

However, inconsistency and variation in the application of the rules was not always due to differences in diagnosis and prognosis but were sometime due to variation in practice across hospitals or between clinicians.

> *“There are two main hospitals in [place name redacted] and the nurses that deal with it in one hospital, it’s like flattening fog, getting something off them. The ones that work at the other hospital, yeah, they will do you one straight away. And that’s [for patients with] the same disease.”* (benefits advisor 7)

#### 2. Positive impacts alongside a sense of shame and stigma

Patients and carers spoke about the positive impact that benefits had on their ability to cover living costs, including paying for travel to appointments, equipment, therapy, medication and housing costs, and for covering other costs including making memories and funeral costs:

> *“it’s certainly been positive it’s…taken a lot of pressure off, I mean it really, really has…as much for my wife as for me…it just takes that pressure off.”* (patient 3)

> *“It is very, very welcome…made a hell of a difference…my wheelchair, hospital appointments…medications I buy for myself vitamin supplements and stuff, and then like if I have to go to the hospital, sometimes I have to go to [ place name redacted] and that’s like a £10 ride there and back…Funeral expenses as well, the money gets put aside for funeral expenses because obviously if I go, as someone who’s on benefits, I don’t have the assets or anything like that to support…Also, the positive little things…we went to [theme park name redacted] on the 8th for my nephew’s birthday, I had the taxi fares saved from that money, so I can make memories with my family in the process of everything that’s happening.”* (patient 5)

Many patients and carers also expressed difficult feelings about claiming benefits, including feeling fraudulent or guilty, having to overcome their pride, and talked about negative stereotypes attached to people claiming benefits:

> *“So, I feel a bit fraudulent, and I said that to my nurse and she said ‘no, don’t be silly, you know, you’re entitled to it’.”* (patient 1)

> *“I refused, I didn’t want it…it were like begging for me, you know.”* (patient 4)

> *“I didn’t like doing it…I’m very much of the opinion that you know, we should support ourselves really, shouldn’t rely on the state. I just think we should be responsible for ourselves, I think, really, as much as we can.”* (carer 1)

Healthcare professionals also recognised the real or perceived stigma that patients can face when claiming benefits and how this can act as a barrier for people, preventing them from claiming benefits.

> *“I think people don’t claim because they’re called welfare, they’re called benefits, they’re called special, and in the main, people are incredibly proud, inappropriately proud at times and don’t want to make claims*.” (social worker 3)

Many of the healthcare professionals reflected on the importance of the language they used, being aware of words that might stigmatise and adjusting their language to make discussions about benefits easier or more acceptable.

> *“I would normally ask if they’d like to have a chat about their finances and benefits…to explore whether they’re getting all that they’re entitled to…I don’t normally use the word ‘benefit’.”* (physiotherapist 3)

Some healthcare professionals described patients who did not want to claim benefits because they did not need the financial support and that raising the question of benefits can be quite surprising to some people. For others, a refusal might be due a sense of social responsibility.

> *“I certainly have had, you know, a couple of occasions where people have been absolutely shocked that I’ve asked the question about, ‘you know, are you receiving any benefits and things’…[and they respond] ‘can you not see where we live, don’t we look you know, look well off etc.’ So, I’ve have had that a couple of times, absolutely.”* (nurse 1)

> *“With people who say no, it definitely comes across as a sense of social responsibility. And often if they then come round to it, it’s because they’ve mentioned it to their children, for example, and their children have gone ‘why have you said no to this? You know, you’re entitled to it, just accept the support’.”* (nurse 3)

#### 3. Talking about money, difficulties and dividends

Several professionals spoke about the need for building trust before talking about finances, and that initial consultations may not be the best time for sensitive discussions about finances.

> *“I think with SR1s you know, you have to have that relationship with the patient to be able to have those conversations.”* (social worker 7)

> *“I think a lot of it also is about relationships, about people trusting you, and sometimes in a first assessment you can’t do all of that.”* (doctor 1)

Some professionals described feeling uncomfortable speaking to patients about finances, recognising that these are sensitive topics and acknowledging that they might not want to discuss their own finances if they were a patient.

> *“I wouldn’t want anyone to know about my personal bit, you know? But yeah, it’s very difficult to ask those questions.”* (social worker 4)

> *“Why do I feel uncomfortable with it? I don’t know. I just feel like…it is quite an intimate thing…feels like quite an accusatory thing, you know…there is something that just feels sort of uncomfortable when trying to explore that, I’ll always tread very gently…I will go there, but I’m always just slightly wary of how I come at that.”* (physiotherapist 3)

Healthcare professionals described the financial impact that serious illnesses can have on high-income patients as well, particularly those of working age, and the sensitivities of discussing finances especially in front of family members.

> *“It was very challenging trying to talk to families that are actually sort of high-income earners, suddenly facing transplant and not having an income, and yeah very difficult. So, I’ve experienced that more recently and I did find it challenging.”* (social worker 12)

Others were more comfortable addressing finances as part of the holistic assessment of patients, and some felt empowered to have those conversations because they were able to offer financial support through the ‘Special Rules’.

> *“I feel completely comfortable about talking about finances because that’s very much part of what we do. You know we need to make sure people have got the support that they need”* (social worker 16)

> *“Asking doesn’t ever feel wildly uncomfortable…especially because…if you know that you’re asking because you’re in a position to offer support, then it’s quite a positive lead in, isn’t it?”* (nurse 3)

Patients and carers also described feeling comfortable with discussions about claiming benefits under the ‘Special Rules’.

> *“I think my nurse explained it all quite thoroughly, she’s brilliant…she kind of sat us down and said ‘right, we can apply for this, all I need to do is fill in this form…we do have to put on it that we expect you to die within 12 months. But what that means is that you get this benefit…’ And she said it’s done under something called special rules because of your illness and you know your short prognosis…some people might say it was too much information, but for me it was perfect.”* (patient 1)

Occupational therapists and physiotherapists explained how discussions about equipment or other costs associated with care at home can be used as a prompt for discussing financial support.

> *“I don’t routinely ask about income benefits when I go on a visit, but…if I identify that someone needs something that we can’t provide, that they might have to pay for, that’s then when I might say, ‘are you receiving any payments that could help to contribute towards that?’”* (occupational therapist 1)

Talking about benefits was also seen by some professionals as a way of introducing other topics such as advance care planning.

> *“I found that the SR1 form provides a really good structure and strategy to then raise other issues naturally, about…advance care planning…the person then raises issues around, where they want to…be buried, cremated, where they want their care and treatment. Particularly, we see…what they want to happen with regard to children etc. So, the SR1 form can actually, you know, provide a really good way of having a wider conversation about current social and financial issues.”* (social worker 3)

Many healthcare professionals had a good knowledge of the ‘Special Rules’, how to process SR1 forms, and of the value of payments. Those less knowledgeable about the value of benefits, tended to underestimate their worth. Some explained how a better awareness of the value of these benefits would make them more likely to prioritise completing SR1 forms.

> *“I think it might be like one taxi fare and my best guess, you know, like it might be really something sort of quite small, you know, like, I don’t know, like…£20 or £30 a week or something”* (physiotherapist 3)

> *“This morning, we were all like, oh God, it’s that much money…then you want it for all your patients, so it becomes more of a priority because you know the difference it will make.”* (nurse 9)

#### 4. Everybody’s – yet nobody’s - responsibility

Healthcare professionals differed in their opinions about who should take responsibility for speaking to patients about benefits and eligibility under the ‘Special Rules’ and indicated that this could lead to a lack of ownership. Some doctors, nurses and social workers saw it as a key part of their role, but others were more reluctant and described the challenges of competing clinical priorities and a lack of knowledge about benefits.

> *“I sometimes find myself no longer a social worker, but a welfare benefits officer, and it’s growing every day. That money is very important for people to have, especially when they’ve gone through such serious life changes. So, I stand in as an advocate, but I have limited knowledge, limited experience.”* (social worker 4)

> *“I think you know you are there to look after the patient’s medical problems aren’t you?…I don’t think you should sort of feel that you’re letting patients down by not discussing benefits…I don’t think that you should be the social worker in the clinic, but you need a process of how to address it maybe outside clinic, I don’t know.”* (doctor 7)

> *“Because its everyone’s responsibility, it ends up being no one’s responsibility.”* (doctor 2)

A reoccurring theme was that for some clinical specialities, particularly those dealing with a lot of uncertainty around the prognosis and survival of their patients, there was a feeling that discussing access to benefits under the ‘Special Rules’ was a job for the specialist palliative care team.

> *“It’s hard, particularly for a CF team or a liver team or a kidney team to actually, I think come to terms with failure…that’s not the right phrase, but I think it is about they want to cure people…it’s someone else’s job to talk about death and dying…we often don’t have the conversations.”* (social worker 3)

Doctors, nurses, physiotherapists and occupational therapists frequently described how support from a dedicated social worker or benefits advisor meant that they felt able to speak to patients about access to benefits under the ‘Special Rules’ and then refer the patient on to someone “*better placed to give the right advice*” (nurse 6). They described the important role of benefits advisors in delivering training, and benefits advisors described how they help to educate clinical colleagues, including by developing their own written guidance.

> *“The SR1 document is very complicated to read…that’s why we made our own real simple SR1 clinical guidance that we send GPs…By giving them the information when we’re requesting [the SR1]…we’re educating them without them knowing we’re educating them”.* (benefits advisor 4)

Clinicians described a ‘postcode lottery’ in the availability of social workers and benefits advisors, citing a lack of access to these members of the team with specialist knowledge of benefits as a major barrier to supporting patients.

> *“The big barrier we have is we don’t have a social worker”* (doctor 2)

A reflection that was shared by several doctors and other healthcare professions, was that while the support of a dedicated social worker or benefits advisor was greatly appreciated, it potentially leads to a deskilling of other members of the team.

> “I’ve been very lucky that I’ve been in places that are really well supported…from a social work kind of point of view, which is fabulous for the patients. But I think it’s probably meant that I’ve been quite ignorant of a lot of this.” (doctor 10)

#### 5. Sticking points in the system

Social workers shared accounts of their interactions with the Department for Work and Pensions (the government organisation which processes benefit claims), with many reflecting positively on the speed at which claims are processed and payments are made under the ‘Special Rules’. Others described challenges in the system, including delays due to missing paperwork, perceived unfairness in the way that decisions are made, and a lack of response to queries.

> *“Occasionally I might have a question in relation to sending an SR1 with regards an individual, but you never get any response.”* (benefits advisor 3)

> *“It’s the way it’s administered and the completely random decisions that you see, between somebody who’s perhaps working, walking, going to a gym, on the upper rate, and then somebody waiting for a lung transplant on the standard rate.”* (social worker 3)

Several social workers and benefits advisors highlighted an inconsistency in the way that applications for different disability benefits are made and felt that the method for the working age disability benefit Personal Independence Payment (PIP) – a phone call that can be made on behalf of the patient - was easier than the paper form needed for the equivalent pension age disability benefit Attendance Allowance (AA) (applications for AA can now also be made using an online form).

However, several healthcare professionals highlighted the insensitivity of the automated message that patients hear when they call the DWP to make a claim for PIP and explained how they helped patients to navigate this.

> *“I mean that [PIP] is just a phone call and it’s very easy…I think the only time it becomes an issue [is] if…they’re a little bit in denial about prognosis…because the DWP [ask] ‘has a healthcare professional told you you’ve got 12 months or less to live?’ which is a very, very different question to ‘would you be surprised if your patient died within the next 12 months?’ Completely different question, completely different emphasis…And it’s quite a harsh question, and sometimes I just say to them, it’s a crass question, just press 3 and you’ll go through.”* (benefits advisor 1)

Benefits advisors described the process of completing and submitting SR1 forms as often involving several members of the team. Sometimes this was to ensure payment for completing forms which is available as an incentive only for doctors.

> *“We kinda do it slightly differently just so that we can get paid. So, our CNSs who actually see the patient will technically complete the paperwork, but then it’s sent to our consultants to be signed because that’s the only way we can actually get paid…for completing the SR1, and then it then goes back into the system for our admin to send off.”* (benefits advisor 1)

The lack of access to shared records was flagged as a potential barrier in the process, since there is no consistent way for healthcare professionals to check whether an SR1 has been completed for a patient.

> *“I think there might be an assumption that it’s already been done as well. It’s hard to check and often patients don’t know themselves what’s been [done], you know…It’s hard for us to know, or to find out if that’s already been done. We don’t really have access to any of that information”* (nurse 7)

One doctor described how since taking part in this study, they had linked the completion and recording of SR1 forms to the completion of advance care plans.

> *“Already this morning we identified three people we would have missed, it’s just that simple change by linking it to [an advance care plan]”* (doctor 2)

Another suggestion was to link completion and recording of SR1 forms to discharge planning.

> *“There’s something about, not necessarily the point of admission to hospital, but on the point of doing discharge planning…it being a standard add-in tab…[for] the nurse or the doctor or whoever is the person that’s doing…[the] checklist for discharge.”* (physiotherapist 3)

## Discussion

This qualitative study provides new evidence on the barriers and facilitators to making benefit claims under the ‘Special Rules for End of Life’ from the perspectives of patients, carers and healthcare professionals. Collectively the themes reveal a number of challenges that may contribute to people not taking up this financial support, including prognosis uncertainty, stigma, professional discomfort talking about finances, a lack of clarity regarding whose responsibility it is to support patients to access this support, and pain points in the system. Discussions about the ‘Special Rules’ also were seen as a core part of holistic care, a positive way to offer support and a gateway to other discussions about end-of-life care preferences and decisions. Overall, the findings point towards several recommendations for system and service level improvement.

Despite the clear material benefits of financial support, experiences of shame and stigma were recognised as significant barriers to claiming among patients, carers and healthcare professionals in this study. Participants frequently described feelings of guilt, fraudulence and threats to personal pride, reflecting wider evidence that claiming benefits can destabilise moral identity and generate self-stigma even in contexts of clear eligibility.^37–39^ Qualitative research with long-term sickness benefit recipients has shown how individuals engage in extensive narrative work to resist stigmatised identities, emphasising past productivity and deservingness in order to distance themselves from pejorative welfare stereotypes.^37^ Quantitative evidence further demonstrates that such stigma is widespread and consequential. A nationally representative study found that over one-quarter of respondents reported stigma-related reasons that would make them less likely to claim benefits, with both personal shame and anticipated judgement from others acting as deterrents.^40^

Our findings echo this distinction between self-stigma and perceived stigmatisation, particularly where healthcare professionals discussed patients who rejected benefits on grounds of social responsibility. Importantly, stigma should be understood not only at the level of social interactions between people and services but also as the product of the political and economic context, whereby institutional stigma is built in to systems and feeds into perceptions and experiences.^41, 42^ Healthcare professionals’ awareness of the sensitivities and importance of their own linguistic framing surrounding benefits reflects a recognition (implicit or explicit) that stigma is socially produced and institutionally reinforced, rather than solely an individual emotional response.^40, 43^ These findings underscore how stigma operates as a structural and symbolic barrier to benefit take-up, including towards the end of life.

Discussions about access to benefits were closely entangled with uncertainty about prognosis in our data, aligning with wider evidence that prognostic uncertainty is pervasive in advanced illness and challenging to address in clinical encounters.^44, 45^ For some healthcare professionals in our study, prognostic uncertainty was a major barrier to discussing access to benefits under the ‘Special Rules’ particularly for some diagnostic groups. This finding mirrors quantitative evidence that the take-up of benefits in the last year of life varies widely by diagnosis.^22^ Similar tensions around prognosis are described in the advance care planning literature, where uncertainty is cited as a barrier to initiating conversations, driven by ethical concerns about causing distress or undermining hope.^46, 47^ Healthcare professionals in our study described how they managed prognostic uncertainty by avoiding definitive prognostic statements and acknowledging uncertainty in their conversations with patients. This strategy of ‘planning for the worst and hoping for the best’ has been described elsewhere as being employed by both clinicians and patients.^48, 49^ Such approaches have clear benefits but may have unintended consequences where avoidance of explicit discussions around prognosis can reduce patient participation and contribute to delays in planning, highlighting the ethical and practical challenges of managing uncertainty rather than engaging with it directly.^50, 51^

## Implications for policy and practice

The findings from this study indicate several implications for policy and practice, summarised in table 1.

|  |  |
| --- | --- |
| **Policy recommendations** | **1. Consideration should be given to shifting eligibility for benefits under the ‘Special Rules’ away from a prognostic threshold towards diagnosis-based criteria.**<br>Scotland offers an example of this approach.<sup>52</sup> Since 2021, eligibility for accessing benefits under the ‘Special Rules’ in Scotland has been determined by the diagnosis of terminal illness (defined as a progressive disease from which death can reasonably be expected) rather than a prognostic timeframe.<br>Our research has highlighted that many healthcare professionals worry about applying the prognostic criteria, and for some patient groups where diagnosis is highly uncertain, the prognostic criteria can be a major barrier to discussing access to benefits via the ‘Special Rules’ with patients.<br>Recent analysis has highlighted wide variation in the take-up of non-means tested disability benefits in the last year of life by cause of death (ranging from 44% for people dying from liver disease to 90% for people dying from neurodegenerative disease).<sup>22</sup> A shift to diagnostic criteria could reduce ethical and practical barriers for clinicians and mitigate inequalities between diagnostic groups.<br>However, more evidence on the likely impact of such a shift is needed. A quasi-experimental study in England and Wales has indicated an increase in benefit take-up under the ‘Special Rules’ following the widening of the prognostic threshold from a 6 to a 12 month prognosis.<sup>53</sup> A similar approach to evaluating the changes to take-up in Scotland following the shift from a prognosis to a diagnosis-based criteria, would be a good first step for informing potential adoption in other parts of the UK.<br>**1. Future policy changes aimed at streamlining the submission of SR1 forms should recognise the central role played by social workers and benefits advisors in some settings.**<br>Social workers and benefits advisors play a central role in advising patients and clinical colleagues on the ‘Special Rules’ and often support the completion of SR1 forms including printing and submitting forms. Where social workers and benefits advisors are not available, this has been highlighted by other healthcare professionals as a major barrier to supporting patients to access support via the |
|  | 'Special Rules'.<br>Future policies that overlook the role of social workers and benefits advisor may risk limiting support. For example, moves towards digitally completed SR1 forms which require clinical personnel to complete and submit SR1 forms directly online, could unintentionally reduce access if other submission routes via paper and email are discontinued.<br>**2. Greater standardisation in benefit application processes is needed.**<br>Variation in application processes (currently applications for PIP under the 'Special Rules' can be made by phone, and applications for AA can be made online or by post), potentially introduce inequalities in access. Standardising a broad range of application routes could support more equitable access.<br>In addition, attention must be paid to automated telephone prompts which can be perceived as insensitive by patients. For example, the PIP application automated prompt that references a life expectancy of twelve months or less was highlighted by healthcare professionals as in need of revision. |
| **Practice recommendations** | **3. Integrate completion of SR1 forms with existing processes such as advance care planning, Electronic Palliative Care Coordination Systems (EPaCCS), hospital discharge planning and GP palliative care registers.**<br>At a system level, there is scope to strengthen professional ownership for completing SR1 forms by integrating completion with existing processes and shared electronic records, enabling teams to identify whether an SR1 has been completed and by whom.<br>**4. Ongoing education for healthcare professionals to increase knowledge and confidence in supporting patients to access benefits via the 'Special Rules' is needed.**<br>Education should include information on:<br>• the monetary value of benefits, which may help to motivate healthcare professionals to have discussions about financial support;<br>• the department for Work and Pensions guidance which presents the 12-month prognostic criteria as a guideline, and clarifies that there are no penalties for patients or healthcare professionals if a patient lives beyond this timeframe;<br>• how to speak to patients about financial issues, how to use language that reduces the stigma associated with claiming benefits, and how to frame discussions about benefits as part of holistic support. |
|  | Informed by the findings from this study, we have developed a 'bite-sized' training session, delivered by SM to 426 healthcare professionals in 26 sessions between July and December 2025. A video of the training is available here: https://www.kcl.ac.uk/research/the-take-up-study |

## Strengths and limitations

A key strength of this study was the inclusion of a wide range of professionals from across health and social care, allowing for broad exploration of different experiences across the system. While not designed as action research, the focus groups with healthcare professionals appeared to raise awareness of benefits entitlements and processes, representing an unintended but positive practice-based impact of the research. An important reflection is that the researcher who facilitated the focus groups (SM) had an existing professional relationship with some healthcare professional participants, which may have influenced group dynamics or contributions, although this also enabled richer discussion through established trust.

Despite robust recruitment strategies, we did not recruit as many patient and carer participants as we wanted to. We were unable to recruit through general practice and did not include any participants who did not speak English, despite having resources to use live interpretation services. Offering reimbursement to patients and carers may have improved participation and inclusivity and should be considered in future studies.^54^ The relatively small number of patients and carers included in this study limits the diversity of perspectives captured, particularly those of under-represented groups such as those from Black ethnic groups, those who did not speak English well, and those who were not in touch with hospital or hospice services.

## Conclusion

This study highlights how access to disability benefits under the ‘Special Rules for End of Life’ is shaped by system design, and by relational and ethical factors embedded in clinical practice. While the material value of financial support at the end of life was widely recognised, stigma, prognostic uncertainty and ambiguity around professional roles in supporting patients to access benefits via the ‘Special Rules’ were barriers to making claims. Discussions about benefits were often bound up with difficult conversations about prognosis, creating particular challenges for those living with conditions that have uncertain trajectories. At the same time, where handled sensitively, conversations about benefits were seen as a meaningful way to offer support and could be a gateway to other discussions about planning and care preferences.

Moving from prognosis-based to diagnosis-based eligibility criteria could help to widen access to benefits via the ‘Special Rules’ and reduce inequalities between diagnostic groups, with Scotland providing a useful model for this. Integrating SR1 completion into existing systems such as hospital discharge planning and Electronic Palliative Care Coordination Systems (EPaCCS), and improving professional education particularly around the value of benefits, eligibility, communication, and stigma, could strengthen take-up and ensure patients receive timely financial support as part of holistic care.

## Data Availability

All data produced in the present study are available upon reasonable request to the authors.

